# Percutaneous rheolytic thrombectomy of thrombosed arteriovenous dialysis access using the AngioJet catheter- A Systematic Review

**DOI:** 10.1101/2024.05.24.24307920

**Authors:** Quinncy Lee, Lap Hin Ho Dominic, Jun Jie Ng, Andrew MTL Choong

**Affiliations:** SingVaSC, Singapore Vascular Collaborative, Singapore; The School of Medicine, Medical Sciences, and Nutrition, University of Aberdeen, Aberdeen, United Kingdom; Faculty of Infectious and Tropical Diseases, London School of Hygiene and Tropical Medicine, London, United Kingdom; School of Nursing and Health Studies, Hong Kong Metropolitan University, Hong Kong, China; Cardiovascular Research Institute, National University Heart Centre Singapore; Department of Surgery, Yong Loo Lin School of Medicine, National University of Singapore; Division of Vascular Surgery, National University Heart Centre, Singapore; Asian Aortic and Vascular Centre, Mount Elizabeth Hospital, Singapore

**Keywords:** Arteriovenous fistula, Arteriovenous graft, Endovascular intervention, Haemodialysis, Rheolytic thrombectomy

## Abstract

**Introduction:** The endovascular removal of thrombi in occluded arteriovenous (AV) access sites has been increasingly accepted by surgeons as an alternative to surgical thrombectomy and revision [1,2]. This systematic review documents the experiences and outcomes of using the AngioJet Rheolytic Thrombectomy Device for salvaging thrombosed arteriovenous fistulas (AVFs) or grafts (AVGs).

**Methods:** Electronic databases, such as PubMed, Embase, and the Cochrane Library, were searched from their establishment until May 2024. Initially, 549 articles were reviewed for potential inclusion, and only 10 studies fulfilled our inclusion criteria. Our final dataset included 771 patients who underwent 996 thrombectomies to treat 338 thrombosed AVFs and 457 AVGs.

**Results:** The demographics, technical and procedural success, patency rates, and complications were evaluated to examine the effectiveness and safety of AngioJet thrombectomy. The mean primary patency rates at 3, 6, and 12 months were 64.09 ± 10.12, 50.36 ± 11.73, and 40.81 ± 15.13 (p < 0.05). The mean assisted primary patency rates at 3, 6, and 12 months were 75.37 ± 13.98, 59.58 ± 14.26 and 40.88 ± 18.81 (p < 0.05). Finally, the mean secondary patency rates at 3, 6, and 12 months were 84.03 ± 8.38, 77.93 ± 9.07, and 65.81 ± 12.12 (p < 0.05). A total of 92 complications were recorded; most were minor complications, thereby being transient and self-limiting. Additionally, 5 total deaths were reported; however, all were not considered related to the AngioJet device.

**Discussion:** Our study deemed devices such as AngioJet efficacious in performing pharmacomechanical thrombectomies, as promising results in terms of safeness and effectiveness to re-establish patency in occluded AVFs and AVGs have been reported.

## Introduction

Chronic kidney disease (CKD) is a significant public health concern, especially in aging populations. The prevalence of CKD inevitably increases with biological age [3]. Subsequently, the demands for treatment, such as haemodialysis, peritoneal dialysis, and kidney transplantation, increase.

Haemodialysis is a temporary treatment for patients awaiting kidney transplants and a permanent treatment for end-stage renal disease (ESRD) patients [4]. In the case of regular haemodialysis, arteriovenous (AV) access is created before the initiation of haemodialysis. There are three main types of AV access: arteriovenous fistula (AVF), arteriovenous graft (AVG), and central venous catheter (CVC) [5].

AVF is the most preferred dialysis access due to the low incidence of associated morbidity and mortality. In the case that the sites for AVF are unavailable, e.g., if patients do not have adequate native veins for creating a fistula, AVG is alternatively used. Central venous catheterisation is not preferred as it entails a high risk of morbidity and mortality due to complications such as catheter-related bloodstream infections (CRBSI) [6–9].

Thrombosis is a common AV access-related complication leading to the dysfunction and failure of AVF and AVG, ultimately causing haemodialysis failure (e.g., delayed dialysis or under-dialysis) [10–11]. Thrombotic occlusions can occur either immediately after AVF or AVG creation or later during follow-ups. The treatment of thrombosis involves removing the thrombus to maintain the functionality of the AV access and restoring the adequate blood flow for haemodialysis.

A major cause of thrombosis is venous outflow stenosis. Additionally, factors such as hypercoagulability, hypotension, inflow, or puncture site stenosis contribute to the risk of thrombosis [12]. Thrombotic occlusions should be treated immediately, as they result in high morbidity and repeated hospitalisation. Additionally, thrombosis-related deaths occur due to cerebrovascular accidents (CVA), myocardial infarction (MI) or stroke [13]. Traditionally, thrombosed AV accesses are treated with surgical interventions. Currently, various interventions are applied, such as aspiration, balloon-assisted, mechanical thrombectomy, or pharmacological thrombolysis [14]. However, there is no existing literature or standardised guidelines indicating which procedure is the most effective for AVF or AVG failure.

This systematic review aims to analyse the existing studies on pharmacomechanical thrombectomy using the AngioJet device in patients with thrombosed AVF or AVG. The complication, patency, and success rates were evaluated to determine the effectiveness of pharmacomechanical thrombectomy compared to other interventions. Furthermore, factors that affect the patency and success rates were identified and discussed in detail.

## METHODOLOGY

### Materials and methods

This systematic review follows the guidelines of Preferred Reporting Items for Systematic Reviews and Meta-Analyses (PRISMA) [15] and Meta-Analyses and Systematic Reviews of Observational Studies (MOOSE) [16].

### Search strategy

The online databases Medline, Embase, and Cochrane were searched; the time frame for the searches was from the date of establishment to May 2024. The following medical subject heading (MeSH) terms were searched: ‘access’, ‘AngioJet’, ‘fistula’, ‘graft’, ‘haemodialysis’, ‘mechanical’, ‘pharmacomechanical’, ‘shunt’ and ‘thrombectomy’. The detailed search strategies for MEDLINE are shown in Supplementary Table 1. Reference lists from comparative or observational studies were cross matched with the search results to ensure that the search terms were broad enough and included all relevant studies.

### Publication selection

For a study to be included in the systematic review, the following inclusion criteria had to be met: 1) study population aged 18 to 85 years; 2) study of patients with thrombosed arteriovenous fistula or graft 3) who were undergoing endovascular declotting and 4) using a mechanical thrombectomy device (AngioJet) as a part of their treatment. Studies of patients with severe cardiopulmonary diseases (e.g., atrial septal defects), non-English articles without a translation, single or small case reports, animal vein thrombosis models, and conference publications were excluded.

Only the study by Yang et al. was included; the studies by Hossain et al. [17] and Wen et al. [18] were excluded from our systematic review, although they fulfilled the inclusion criteria. This is due to significant similarities in the reported data, which suggest that Hossain et al. and Wen et al. used the same participants as Yang et al.

### Data extraction

First, two authors independently screened the studies for inclusion based on the abstracts and titles. Studies that were included based on this initial screening were then reviewed in the second stage. Data extraction followed, and a third reviewer verified the information collected. Any conflicts among reviewers were resolved by consensus. Final decisions on inclusion in the study were made by all reviewers.

### Outcomes

Success and patency outcomes were defined based on the reporting standards of the Society of Interventional Radiology (SIR). Anatomic success was defined as restoration of flow combined with less than 30% maximal residual diameter stenosis after treatment [19]. Clinical success was defined as the completion of at least one haemodialysis session after treatment [20]. Technical success was defined as the restoration of blood flow, combined with residual stenosis of less than 30% [21].

Primary patency was defined as the interval from the first AngioJet thrombectomy to the next thrombosis or reintervention. Assisted primary patency was defined as the interval from the first AngioJet thrombectomy to the next thrombosis or reintervention, including prevention methods or treatment for AVF or AVG failure. Secondary patency was defined as the interval from the first AngioJet thrombectomy until the fistula or graft required surgical declotting and revision [21].

### Statistical analysis

Since only a small number of studies were included, weighted pooling of data, such as a meta-analysis of proportions of means, was not performed because of the risk of bias due to sampling errors. Furthermore, there was significant heterogeneity in the reporting methods of the included studies. These included different measurements of pre-dialysis comorbidities, different time intervals from initial diagnosis of thrombosed AVF or AVG to endovascular intervention, different post-dialysis success rates, and different durations from initial treatment to follow-up for patency rates.

## RESULTS

### Study Characteristics

In the initial literature search, 549 articles were identified (232 from MEDLINE, 294 from EMBASE, and 23 from the Cochrane database). The articles were initially screened for inclusion by abstract and title. Next, the full text of each selected article was reviewed and evaluated in detail for further exclusion. In the end, the study design included 3 prospective and 7 retrospective studies. The study identification, selection process, and reasons for exclusion are illustrated in the PRIMSA flowchart (Figure 1).

**Figure 1:**
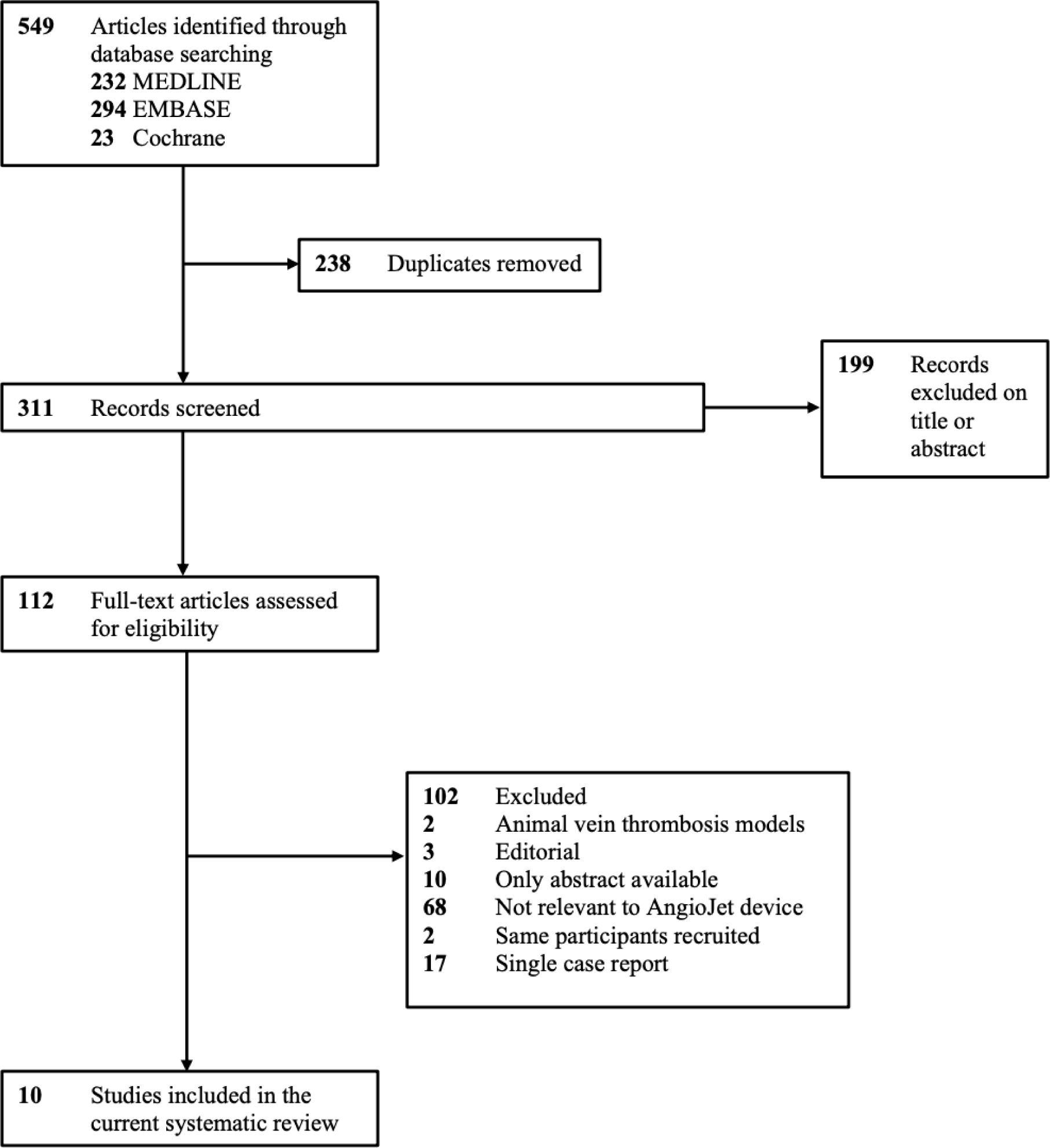
PRISMA flow diagram for study selection process

Of the studies, 2 were conducted in the United States and 1 each in Belgium, Italy, Korea, the Netherlands, Spain, Taiwan, Turkey, and the United Kingdom. Of the 10 studies, 3 were prospective and 7 were retrospective.

### Patient selection

A total of 771 patients underwent 996 AngioJet thrombectomies for treating 338 AVFs and 457 AVGs. In 2 studies, patients with only AVF were included; 3 studies were on patients with only AVG, and 5 studies included patients with both AVF and AVG.

The patients’ existing comorbidities were not reported in 3 studies [21, 22, 26], and in the remaining 7, they were not reported consistently [23–25, 27–31]. Common comorbidities, such as diabetes mellitus (DM), hypertension (HTN), smoking habits, history of peripheral vascular disease (PVD), hyperlipidaemia (HPLD), and dyslipidaemia (DLP), were reported in 7, 6, 4, 4, 3 and 2 studies, respectively. Body mass index (BMI) was reported heterogeneously in 2 studies [29, 30]; the mean BMI of patients in the AVF and AVG groups [29] and the number of individuals with a BMI greater than 30 were also provided [30]. These results are summarised in Supplementary Table 2.

One study also reported the known hypercoagulability, surgery in the past 30 days, current malignancy, and contradiction to thrombolytic agents [24]. The platelet count and prothrombin time (INR) of patients were recorded in one study [30]: 53.3% (n = 32) and 46.7% (n = 28) of patients had a platelet count of 150,000–300,000 x μL and 300,00–450,000 x μL, respectively. The INR was reported to be 0.8–1.2 and >1.2 for 93.3% (n = 56) and 6.7% (n = 4) of patients, respectively.

Furthermore, the pre- and post-dialysis serum potassium levels were measured. During pre-dialysis, routine blood analyses were performed to ensure that the serum potassium was in the normal range, and after dialysis, routine blood analyses were performed again to ensure no significant increase in serum potassium [21].

### Types of haemodialysis vascular access

The details of haemodialysis vascular access are also summarised in Supplementary Table 2. When reported, the most common sites of AVF creation were brachiocephalic fistulas (n = 50) [21, 22, 26, 29], followed by radiocephalic fistulas (n = 47) [21, 22, 26, 29]. A fraction of 56.0% [26], 62.5% [22], and 74.6% [23] of AVFs were created on the non-dominant arm. The most common sites of AVG creation were brachioaxillary grafts (n = 55) [21, 27, 30], with 67.8% [22], 69.1% [27] and 89.7% [28] of AVGs created on the non-dominant arm.

Although 2 studies did not report the type of catheter used [28, 31], a variety of AngioJet catheters were used in the remaining studies. More than a decade ago, earlier models (i.e., F-105) were used in patients receiving a thrombectomy [23]. Newer generations and advanced models of AngioJet catheters, such as AVX [23–24, 26, 29], DVX [21, 24], Solent-Proxi [29, 30], Spiroflex [24] and Xpeedior [24], were used in later studies. In 3 studies, more than one type of catheter was used for occluding thromboses in AV access sites [23, 24, 29]. The heterogeneity of catheter use was probably due to the availability of catheters, the clinical conditions of the patient, and the practitioner’s experience [28].

### Number of stenoses requiring angioplasty and/or stenting

Following pharmacomechanical thrombectomies, patients with stenotic lesions were treated by either percutaneous transluminal angioplasty (PTA), stenting, or surgical intervention. In those reported, 82% [24], 91.7% [30], 98% [31], and 100% of PTAs were carried out following AngioJet thrombectomy [21, 27–28]. Moreover, 41% (n = 7) [27], 42% (n = 30) [24] and 53% (n = 34) [21] of the patient population from 3 studies required stent insertion to treat angioplasty-resistant stenoses, as summarised in Supplementary Table 3.

In one study, after PTA, primary failure (PF) occurred only in 10% of the patient population, with successful dialysis use in the remaining 90% [31]. We should also mention that patients with a history of AV access site dysfunction or thrombosis and prior interventions received a higher median number of stenoses than those without a history (median 4, IQR 3–4 vs median 3, IQR 3–4 (p = 0.07)) [22].

Following PTA, in one study, additional PTAs, venous stent placement and surgical interventions were used to remove residual stenosis in the AVF and AVG patient groups [29]. Additional PTAs were required in 88.9% of the AVF and 55.4% of the AVG groups, venous stenting in 3.7% of the AVF and 10.8% of the AVG groups and surgical intervention in 18.5% of the AVF and 13.8% of the AVG group (p = 0.391) [29]. Additionally, after angioplasty and/or stenting, 2 studies reported additional interventions: 10% of procedures required restenosis of the treated vessel within a 3-month follow-up period in one study [24], and 24% of cases required multiple stenosis in another study [23]. Moreover, in two separate studies, resistant stenosis was treated via high-pressure balloon angioplasty (1%; p < 0.07) [23] and cutting balloon angioplasty (3%; p < 0.20 [23] and 11.4% [27]).

### Complications

A total of 92 complications were reported and summarised in Supplementary Table 4. Most complications were transient and self-limiting and were therefore classified as minor [20]. Minor adverse events included arrhythmia (n < 7), bleeding and haematoma (n = 12), bronchospasm (n = 1), and nausea (n = 2); all these patients recovered without any further intervention.

Major adverse events included distal arterial emboli in 7 patients; those reported were treated via Fogarty thrombectomy (n = 2), catheter-directed thrombolysis (n = 1), or percutaneous thromboaspiration (n = 2). None of the cases resulted in limb amputation, organ dysfunction, or death. Reported vein ruptures (n = 5) were treated with covered stent grafts (n = 4) or by surgical revision and transfusion (n = 1). Arterial anastomosis ruptures (n = 2) and a pseudoaneurysm (PSA) of artery-graft anastomosis (n =1) were surgically corrected. A single case of acute myocardial infarction (AMI) 4 hours post-thrombectomy [21] and one case of vascular dissection [24] were reported.

A total of 5 deaths were recorded [21, 24]. Despite immediate thrombolysis and reperfusion of the myocardium, one patient still passed away 6 hours post-intervention [21]. During initial hospitalisation, 2 deaths arose due to cardiogenic shock and MI [24]. Before the three-month follow-up, 2 patients died of coronary vascular disease and an unknown cause [24]. Nevertheless, the authors did not consider the deaths to be related to the use of the AngioJet device for thrombectomy [21, 24, 25], as the effects of arrhythmia were transient and resolved shortly after inactivation of the AngioJet device [24]. The deaths were suspected to be a result of the existing medical comorbidities of the patients.

### Drug interventions

The use of heparin intraoperatively was reported in 8 studies, with significant heterogeneity in the dosage used [21–23, 26, 28–31]: intravenous injections of 1500 IU [30], 2500 IU [29] and up to 5000 IU [21, 26, 31]; in [28], 29 patients were given 521.8 units of heparin; 70 U/kg of unfractionated heparin were injected intravenously in [22]; in [23], 90 out of 134 patients were given intra-procedural heparin, but the exact dosage was not reported. Other anticoagulants used included apixaban (n = 2), rivaroxaban (n = 3), and warfarin (n = 4) [30].

In AVF patients, the use of postoperative (p = 0.003) anticoagulants was found to be significantly associated with secondary patency. In AVG patients, the use of postoperative anticoagulants was found to be significantly associated with both primary (p = 0.009) and secondary (p = 0.010) patency [29]. Littler et. al reported the use of either aspirin (75 mg/day) or clopidogrel (75mg/day); Ierardi et. al reported the use of 100 mg aspirin, whereas Yang et. al administered aspirin or clopidogrel routinely for 3 days (the dosage was unspecified). However, there were no justification studies on whether aspirin or clopidogrel had a better clinical outcome. Additionally, patients who did not receive anti-platelet or anti-thrombotic drugs were given a low dosage of acetylsalicylic acid to reduce inflammation [29].

### Follow-up duration, success rates, and patency outcomes

Common intervals for follow-up duration were 3, 6, and 12 months. The shortest follow-up duration was immediately after thrombectomy [21, 24], and the longest follow-up duration was > 24 months post-thrombectomy [30].

One study did not measure anatomic, clinical, or technical success [21]. Anatomic success was recorded in only one study (77%) [23]. In [24], only technical success was recorded; clinical and technical success rates were reported separately for AVFs and AVGs [24, 29]. In another study, only overall success was recorded, but not classified [31]. Both clinical and technical success after percutaneous thrombectomy were measured in 5 studies; clinical success ranged from 86% to 97%, and technical success from 93% to 98% [22, 26–28, 30].

The mean primary patency rates at 3, 6, and 12 months were 64.09 ± 10.12, 50.36 ± 11.73 and 40.81 ± 15.13 (p < 0.05), respectively. The mean assisted primary patency rates at 3, 6, and 12 months were 75.37 ± 13.98, 59.58 ± 14.26, and 40.88 ± 18.81 (p < 0.05), respectively. The mean secondary patency rates at 3, 6, and 12 months were 84.03 ± 8.38, 77.93 ± 9.07, and 65.81 ± 12.12 (p < 0.05), respectively. Separate patency rates for AVF and AVG were published [21, 24, 29], and 3-month patency rates were recorded in only one study [24]. A detailed summary of follow-up duration, success, and patency rates are shown in Supplementary Table 5.

### Other factors

The time interval from the initial diagnosis of thrombosis to percutaneous thrombectomy varied greatly among the studies. The shortest time until intervention was 0.4 and 0.6 days after the identification of a thrombosed AVF and AVG [29], whereas the longest time until intervention was 16 days [21]. For the other studies, the time until intervention ranged from 1 to 5 days [22, 26, 27, 30, 31], and most thrombosed AVFs and AVGs were treated ≤ 2 days post-diagnosis [22, 26, 27, 30].

Four studies reported the total procedure time, with a range of 43–127 minutes [23–24, 26, 29]. Two studies reported the length of occlusion for thrombosed AVFs and AVGs; the median length of the thrombosed AVF segment was 12 ± 5 cm [26], and the average length of occlusion for both thrombosed AVF and AVG segments was 24 cm [21]. Moreover, only one study reported the total bleeding amount (51.7 ± 17.6 mL) after intervention [28], the number of repeated AVG thromboses (n = 24) required in less than 1 month and the average time of reintervention after thrombectomy (range: 1–68 days) [27].

## Discussion

Surgical treatment is the gold standard for treating thrombotic occlusion of AV accesses [32–34]; according to the National Kidney Foundation-Kidney Disease Outcomes Quality Initiative (NKF-KDOQI), it is preferred over endovascular interventions although the procedure has significant disadvantages. Surgical thrombectomy has led to large volume blood loss and direct damage to the AV access sites, and it is incapable of detecting additional stenoses [35].

In the 21^st^ century, endovascular interventions have become an increasingly popular alternative to surgery. The AngioJet device creates a Venturi/Bernoulli-effect that fragments and aspirates the clot, preventing clot embolization [36]. Endovascular interventions cause minimal bleeding due to the absence of incisions. In one study, total bleeding was reported to be significantly lower in individuals undergoing AngioJet PMT (51.7± 17.6 mL) than individuals undergoing surgical thrombectomy (72.5 ± 30.7 mL) [28]. This suggests that patients with an identified risk of bleeding can undergo AngioJet thrombectomy as an alternative.

Previous studies mentioned that rapid improvements in endovascular equipment, techniques and skills have led to improved percutaneous rheolytic thrombectomy outcomes when using the AngioJet [31, 37, 38]. Older studies showed that the surgical treatment group had better technical success and primary patency rates than the AngioJet group (78.8% vs 73.2%). Primary patency rates at 1, 2 and 3 months were higher in the surgical group than the AngioJet group (41%, 32% and 26% vs 32%, 21% and 15%, respectively) [25].

The primary patency rates at 6 and 12 months were reported to be higher for the AngioJet group than the surgical group (85% and 67% vs 78% and 55%, respectively, *p* <0.05); primary failure was also lower in the AngioJet group than the surgical group (10% vs 28%, *p* =0.18) [31]. Newer generation and advanced systems, such as the DVX catheter, were reported to be 5-fold more effective than older systems (e.g., F105 catheter) and were more successful for occluded thrombi with greater volume and larger diameters. Three-months after thrombectomy, the primary patency rate was reported to be 46% for DVX and 15% for F105 [27].

The clinical success rates for AngioJet thrombectomy reported in our systematic review (range: 76%–97%) were comparable to other endovascular interventions: 89% for Amplatz thrombectomy [39], 79%–90% for Arrow-Trerotola PTD [40, 41], 85% for hydrolyser catheter [42], 76%–93% for manual thromboaspiration [43–45], 86% for oasis catheter [46], 100% for rotating mini-pigtail catheter [47] and 82%–90% for thrombolysis [48, 49].

The 6-month primary patency rates for AngioJet (range: 39.7%–96.1%) were comparable to other endovascular interventions: 52% for Amplatz thrombectomy [39], 38%–60% for Arrow-Trerotola PTD [40, 41], 50% for hydrolyser catheter [42], 18%–72% for manual thromboaspiration [43, 45], 50% for oasis catheter [46], 47% for rotating mini-pigtail catheter [47] and 71%–81% for thrombolysis [48, 49].

The 12-month primary patency rates for AngioJet (range: 23%–78%) were also comparable to other endovascular interventions: 27% for Amplatz thrombectomy [39], 18% for Arrow-Trerotola PTD [40], 50% for hydrolyser catheter [42] and 64%–70% for thrombolysis [48, 49].

In our systematic review, only two studies conducted routine blood testing [21, 30], including serum potassium levels [21], platelet count and prothrombin time [30]. Patients with high pre-dialysis serum potassium and low dialysate potassium concentrations were found to be associated with a high risk of cardiovascular and all-cause morbidity and mortality [50].

Mean platelet volume (MPV) was proposed to predict vascular access (VA) dysfunction in haemodialysis patients. Significant association was found between MPV and VA, OR 1.52 (1.13–2.07), *p* <0.001 [51]. Incidence of VA dysfunction was higher in patients with the highest MPV: 59%, 34%, 27% and 18% for patients in the fourth, third, second and first quartile of MPVs, respectively (*p* =0.001) [51]. Short prothrombin time (INR) was confirmed to lead to higher rates of unsuccessful use of AV access site for haemodialysis (FUHD) [52]. Area under curve (AUC) for prothrombin time and FUHD was 0.645 (0.95 CI, 0.494–0.795) [52].

The Peripheral Use of AngioJet Rheolytic Thrombectomy with a Variety of Catheter Lengths (PEARL) registry has vaguely mentioned the association between the AngioJet device and the development of acute kidney injury (AKI) [53]. AKI is the result of intravascular haemolysis and haematuria due to high-pressure jet sprays [25, 54, 55]. Recently, it has been a complication increasingly reported by AngioJet thrombectomy patients.

Bleeding and haematoma were reported in six studies [21, 24, 27–28, 30–31]. Escobar et al. found a more than 8-fold increased risk of AKI using the AngioJet (OR 8.2, 0.95 CI 1.98–34.17, *p* =0.004) [56]. Shen et al. reported a 22.8% (*p* =0.013) increased risk of AKI in patients undergoing AngioJet [57]. Intravascular haemolysis was confirmed by post-procedural routine blood tests (such as: lactate dehydrogenase, haemoglobin and haptoglobin serum levels) and haemoglobinuria via urine dipstick [25, 53].

Additionally, plasma haemoglobin levels cause the release of intracellular potassium, resulting in hyperkalaemia [25], one of the causes of arrhythmia as revealed in transient electrocardiogram disturbances (ST depression, T wave inversion, peaked T waves, bradycardia and bigeminy) in AngioJet patients. These signs lasted up to 3 minutes but did not alter the treatment [21]. Vesely et al. reported that serum potassium levels returned to baseline within 7 days of intervention [25]. The release of adenosine and other by-products of haemolysis can also cause arrhythmias [21]. In future studies, biomarkers from routine blood tests should be included as they can predict the risk of AKI, hyperkalaemia, and adverse cardiac outcomes.

The Florence Appraisal Study of Rheolytic Thrombectomy (FAST) results showed a significant improvement in epicardial coronary flow (*p* <0.01), frame count (*p* <0.01) and myocardial blush (*p* <0.01) in 116 AMI patients after using the AngioJet. The occurrence of in-hospital major adverse cardiac events (MACE), including cardiac death, myocardial infarction, and target vessel revascularization, was also infrequent. MACE was only reported in 8% of the patient population [58]. Hence, deaths due to acute myocardial infarction (AMI) [21, 24] and cardiogenic shock [24] were not considered in some studies due to the use of the AngioJet [21, 24, 25]. In general, postoperative complications, such as adverse cardiac outcomes, haemorrhage, nausea, pseudoaneurysm (PSA), vascular dissection and venous rupture, are common complications of endovascular and vascular procedures, so it was not unusual to see them in patients undergoing AngioJet thrombectomy. Existing medical comorbidities of patients before AngioJet thrombectomy would be one of the most likely causes of the reported deaths.

In our systematic review, percutaneous transluminal angioplasty was performed in nearly all the thrombosed AVFs or AVGs, after removing the thrombus with the AngioJet. This enables any underlying stenosis to be corrected, preventing recurrent thrombosis of the AV access sites. Stenosis is usually initiated by endothelial cell injury, leading to smooth muscle proliferation and neointimal hyperplasia [59–61].

A meta-analysis by Palmer et al. found that antiplatelet agents, such as clopidogrel and aspirin, demonstrated a protective effect from early thrombosis and reduction of patency in AVFs, but the effect on patency in AVGs remains unclear [62]. No association was found between antiplatelet use and the risk of bleeding or haemorrhage [62]. However, the long-term impacts of antiplatelet drugs and primary/assisted primary/secondary patency rates remain unknown. Low-intensity warfarin was not associated with reducing thrombotic events [63]. A randomised trial by Lok et al. showed a 50% reduction in the number of thrombosis events (1.71 vs 3.41, IRR 0.50, 0.95 CI, 0.36–0.72, *p* <0.001) and the number of interventional procedures to maintain the AVG (2.89 vs 4.92, IRR 0.59, 0.95 CI, 0.44–0.78, *p* <0.001) after fish oil supplementation per 1000 access days [64]. This was also confirmed by a smaller-scale study in which the 12-month patency rates improved 5-fold [65], suggesting that supplements can be an alternative to traditional antiplatelet agents.

Multivariate analyses revealed that older autogenous fistulae (*p* =0.045) and previously stented efferent veins (*p* =0.019) were at risk for early re-thrombosis [26]. Older thrombi were also found to be highly adherent to the vessel wall, making them more difficult to remove than new thrombi [53]. Higher clinical success was found in previously untreated patients when the procedure was conducted within 24 hours from the clinical onset of thrombosis [30]. This suggests the importance of early diagnosis and immediate intervention to avoid re-thrombosis of AVF or AVG.

## Limitations

There are several limitations in the present study. First, only 30% of the included studies were prospective and the remaining 70% were retrospective; moreover, majority of the retrospective studies enrolled few patients. Second, drug interventions and AngioJet catheter selection were at the sole discretion of practitioners, which may have induced a selection bias. Third, no statistical comparison was performed among the different techniques for salvaging thrombosed AVFs and AVGs. Fourth, the literature search was limited to articles in English and those translated into English, and the relevant publications in other languages may have been missed. Finally, a meta-analysis was not conducted due to the heterogeneity in data reporting methods.

## Conclusion

Maintenance of AVFs and AVGs for haemodialysis is paramount [66]. The goal of treating thrombosed AVFs or AVGs is to restore and maintain the access function for the longest possible period. Despite difficulties in comparing results across studies due to the heterogeneities in definitions as well as reporting and assessment methods, AngioJet thrombectomy remains effective in removing thrombi and re-establishing blood flow of AVFs or AVGs for haemodialysis. Compared with surgical thrombectomy, minimally invasive endovascular interventions yield lower morbidity, longer-term disability, complication rates, and hospital stay duration. To achieve optimum outcomes, patients with thrombosed AVFs or AVGs should be referred for thrombectomy at the earliest. Prompt treatment and intervention are essential to maintain the success and patency rates and reduce the complication rates of thrombectomy. In future studies, prospective and large-scale randomised controlled trials using standardised reporting methods and follow-up periods are warranted to confirm the true benefits and potential harms of AngioJet.

## Data Availability

All data produced in the present study are available upon reasonable request to the authors

