## Supplementary Table 1 for "Percutaneous rheolytic thrombectomy of thrombosed arteriovenous dialysis access using the AngioJet catheter- A Systematic Review"

| **#** | **Searches** |
| --- | --- |
| 1 | angiojet |
| 2 | rheolytic |
| 3 | pulsed |
| 4 | pharmacomechanical |
| 5 | mechanical |
| 6 | 1 or 2 or 3 or 4 or 5 |
| 7 | thrombectomy |
| 8 | thrombolysis |
| 9 | declotting |
| 10 | 7 or 8 or 9 |
| 11 | fistula |
| 12 | fistulae |
| 13 | graft |
| 14 | grafts |
| 15 | access |
| 16 | shunt |
| 17 | shunts |
| 18 | 11 or 12 or 13 or 14 or 15 or 16 or 17 |
| 19 | haemodialysis |
| 20 | hemodialysis |
| 21 | dialysis |
| 22 | 19 or 20 or 21 |
| 23 | 18 and 22 |
| 24 | 6 and 23 |
