## Supplementary Table 2 for "Percutaneous rheolytic thrombectomy of thrombosed arteriovenous dialysis access using the AngioJet catheter- A Systematic Review"

| **First Author, Year** | | **No. of thrombectomies**  **No. of patients**  **Country of origin**  **Mean/Median age (years)**  **No. of male (% of male)**  **Study design** | **Types of haemodialysis vascular access** | | | | **Comorbidities** |
| --- | --- | --- | --- | --- | --- | --- | --- |
|  |  |  | **AVG** | **Details of AVG** | **AVF** | **Details of AVF** |  |
|  | Adyin, 2020  [31] | 42  96  Turkey  58  n=25 (59%)  Retrospective | - | - | 42 | **Location**  Forearm: 26  Upper arm: 16 | DM: 30  CHF (EF <40%): 8  PVD: 12 |
|  | Drouven, 2020  [29] | 92  60  Netherlands  AVF/AVG: 58.6/59.4  AVF/AVG: n=36 (55%)/ n=20 (74%)  Retrospective | 65 | **Type**  Straight PTFE: 20  PTFE loop: 45 | 27 | **Type**  Radiocephalic: 13  Brachiocephalic: 10  Basilic vein  transposition: 4 | BMI  AVF: 25.9, AVG: 27.9  DM  AVF: 4, AVG: 25  HTN  AVF: 21, AVG: 53 |
|  | Lee, 2020  [28] | 29  62  Korea  69.28 ± 10.17  n=11 (37.9%)  Retrospective | 29 | **Side**  Non-dominant arm: 26    **Location**  Upper arm: 19  Forearm: 10 | - | - | DM: 24  HTN: 20  DLP: 7  Smoking: 7 |
|  | Ierardi, 2020  [30] | 60  60  Italy  52 ± 7.892  n=35 (58%)  Retrospective | 60 | **Type**  Radio-basilic forearm loop: 27  Brachio-basilic arm loop: 14  Brachio-axillary arm loop: 10  Brachio-basilic straight: 9 | - | - | HTN: 45  Smoking: 31  HCh: 35  IHD: 27  DM: 15  BMI >30: 9  COPD: 8 |
|  | Bermudez, 2016  [27] | 149 in 68 grafts  39  Spain  69.9 ± 13.2  n=2 (57.4%)  Retrospective | 68 | **Type**  Brachial axillary: 36  Forearm loop: 5  Femoral loop: 27    **Side**  Left side: 47  Right side: 21 | - | - | HTN: 43  DM: 22  DLP: 26  IC: 12  CVD: 8  PVD: 9  AF: 9 |
|  | Maleux, 2015  [26] | 39  38  Belgium  70.7 ± 13.8  n=24 (63.2%)  Retrospective | - | - | 39 | **Type and location**  **Elbow**  Brachiobasilic: 7  Brachiocephalic: 10  Brachial artery-median  vein: 1  Radiobasilic: 4  Radiocephalic: 3  Radial artery-median vein: 1  **Wrist**  Radiobasilic: 1  Radiocephalic: 12  **Side**  Right: 16  Left: 23 | - |
|  | Simoni, 2012  [24] | 72  72  USA  62.75  n=38 (53%)  Prospective | 44 | **Location**  Upper extremity: 44 | 28 | **Location**  Upper extremity: 28 | Known hypercoagulability: 6  Surgery in past 30 days: 12  Contradiction to thrombolytic agents: 1  Smoking: 36  DM: 17  Renal insufficiency: 5  Renal transplant: 1  Current malignancy: 5  HTN: 25  HPLD: 14  CAD: 18  CVA/stroke: 9  PAD: 5 |
|  | Yang, 2012  [23] | 134  109  Taiwan  61.0 ± 14.0  n=62 (57%)  Retrospective | - | - | 134 | **Fistula location**  Upper arm: 36  **Side of fistula**  Right-side: 34 | DM: 31  HTN: 42  Smoking: 22  HPLD: 8 |
|  | Litter, 2009  [21] | 64  48  UK  59  n=24 (54%)  Prospective | 20  (in 14  patients) | **Graft type**  Brachioaxillary: 9  Brachiocephalic loop: 3  Brachiobasilic: 1  Femoro-femoral: 1 | 44  (in 34  patients) | **Fistula type**  Brachiocephalic: 19  Radiocephalic: 8  Transposed brachiobasilic: 7 | - |
|  | Kakkos, 2008  [22] | 285  187  USA  AVF/AVG: 63/63  AVF/AVG: n=17 (71%)/n=108 (41%)  Prospective | 261 | **Type**  Loop: 104  Straight: 157    **Location**  Forearm: 69  Upper arm: 184  Thigh: 8    **Side**  Left: 177  Right: 84 | 24 | **Type**  Radiocephalic: 11  Brachiocephalic: 11  Transposed brachiobasilic: 2  **Side**  Left: 15  Right: 9 | - |
