## Supplementary Table 3 for "Percutaneous rheolytic thrombectomy of thrombosed arteriovenous dialysis access using the AngioJet catheter- A Systematic Review"

| **First Author, Year** | **Type of catheter** | **Time interval since initial diagnosis of thrombosis** | **History of intervention for dysfunction or thrombotic events** | **Previous pVA** | **Number of stenoses treated** | **Other factors** |
| --- | --- | --- | --- | --- | --- | --- |
| Adyin, 2020 [31] | NR | Between 1 to 5 days | NR | NR | Angioplasty: n=41 (98%)   a) PF: n=4 (10%) b) Success: n=37 (90%) | NR |
| Drouven, 2020  [29] | AVX or Solent-Proxi | AVF: 0.4 days  AVG: 0.6 days | AVF: 9, AVG: 32 | NR | Number of stenoses: NR  Angioplasty  PTA arterial anastomosis: AVG: n=6 (9.2%)  PTA venous anastomosis: AVG: n=14 (21.5%)  Venous stent  AVF: n=1 (3.7%), AVG: n=7 (10.8%)  Surgical intervention  AVF: n=5 (18.5%), AVG: n=9 (13.8%) | NR |
| Lee, 2020 [28] | NR | NR | NR | Age of existing graft  (months):  23.80 ± 3.039 | NR* | Total procedure time/min:  56.3 ± 13.3  AngioJet working/s: 146.5  Thrombolysis/min: 16.1  Total bleeding amount/mL: 51.7 ± 17.6 |
| Ierardi, 2020  [30] | Solent-Proxi | Less than 1 day:  n= 32 (53.3%)  Between 1 to 3 days:  n=18 (30%)  More than 3 days:  n=10 (16.7%) | Fogarty: n=5 (8.3%) prior first AngioJet thrombectomy  Surgical revision of arterial anastomosis: n=3 (5%) | previous aVA: 2.9  previous CVC: 3  pVA age (months):  9.00 ± 3.039 | Number of stenoses: n=55 (91.7%)  Location of stenoses  Graft vein anastomosis: 23  Outflow: 11  Central stenosis: 2  Artery-graft anastomosis: 4  Graft-vein, artery-graft anastomosis: 8  Graft-vein anastomosis, outflow: 7 | NR |
| Bermudez, 2016  [27] | NR | ≤ 2 days: n=127 (85%)  > 2 days: n=22 (15%) | Previous thrombectomies:  n=0.98 (±1.21) | n=3.4 (previous AVF  or AVG)  Time from graft implantation/months: 22.8 (±18.7) | Number of stenosis: NR  Location of stenosis:   1) Venous anastomosis (72.6%) 2) Arterial anastomosis (2%) 3) Body of graft (11.8%) 4) Both anastomoses (15.4%) 5) Additional central vein stenosis (n=3)   Subclavian vein (n=2)  Iliac vein (n=1) | Repeated AVG thrombosis  in less than 1 month: n=24    Median number of  procedures per patient: 2.2    Average time of  reintervention: 1-68 days |
| Maleux, 2015  [26] | AVX | Within 1 day | NR | n=9 (previous  interventions for stenosis)  n=5 (fistula with a stent inserted previously)  Mean age of fistula: 3.4 years (range: 0.1-11.7) | Angioplasty: at least one performed for all rheolytic thrombectomy procedures  Stent  n=16 (11 Maris, 3 Epic and 2 Zilver)  Location of stent:   1) Cephalic arch (n=3) 2) Upper third of cephalic vein (n=2) 3) Mid third of the cephalic vein (n=5) 4) Distal third of cephalic vein (n=1) 5) Proximal third of transposed basilic vein (n=2) 6) Mid third of the transposed basilic vein (n=2) 7) Distal third of forearm cephalic vein (n=1) | Median length of  thrombosed segment:  12 ± 5 cm    Total procedure time/min:  78.4 ± 31.8  (range: 25-160) |
| Simoni, 2012  [24] | AVX, DVX, Spiroflex  and Xpeedior | Within 14 days | n=7 requiring endovascular intervention in the past 7 days | n=3 for prior  intervention in the same area | Angioplasty: n=59 (82%)  Stent placement: n=30 (42%)  3-month follow-up  n=7 (requiring restenosis of treated vessel) | Mean procedure time/hours  without thrombolysis: 1.4 |
| Yang, 2012 [23] | F-105, AVX | NR | NR | Age of existing fistula (months): 43±49 | Number of stenosis: NR  Multiple stenosis: n=32 (24%)  Resistant stenosis: n=5 (4%)   a) Cutting balloon n=4 (3%) b) High-pressure balloon n=1 (1%) | Procedure time/min:  88 ± 39 (p<0.001)  Thrombosis > 2 days:  n=12 (9%) |
| Litter, 2009 [21] | DVX | Average time: 4 days  (Range: 1-16 days) | - | - | Angioplasty: 100% (n=64)  Stent: 53% (n=34) | Average length of  occlusion treated: 24cm |
| Kakkos, 2008  [22] | AVX | 2 days for AVF and AVG | AVF: n=13  AVG: n=144 | NR | AV access  Number of stenoses:  Previously treated: median 4, IQR 3-4  Non-treated: median 3, IQR 3-4  AVF: median 2, IQR 2-3  AVG: median 4, IQR 3-4 | NR |

*: exact number of stenoses was not reported
