## Supplementary Table 4 for "Percutaneous rheolytic thrombectomy of thrombosed arteriovenous dialysis access using the AngioJet catheter- A Systematic Review"

| **No. of complications** | **Included studies** | | | | | | | | | |
| --- | --- | --- | --- | --- | --- | --- | --- | --- | --- | --- |
|  | **Adyin,**  **2020**  [31] | **Drouven, 2020**  [29] | **Lee,**  **2020**  [28] | **Ierardi,**  **2020**  [30] | **Bermudez, 2016**  [27] | **Maleux,**  **2015**  [26] | **Simoni,**  **2012**  [24] | **Yang,**  **2012**  [23] | **Litter,**  **2009**  [21] | **Kakkos**  **2008**  [22] |
| **Reported** | **3** | **NR** | **3** | **4** | **7** | **3** | **31** | **19** | **20** | **2** |
| AMI | - | - | - | - | - | - | - | - | 1 | - |
| Arrhythmia | - | - | - | - | - | - | 1 | - | <6 * | - |
| Arterial Emboli | - | - | - | - | - | 3 | - | 1 | 2 | 1 |
| Arterial anastomosis rupture | - | - | - | 1 | 1 | - | - | - | - | - |
| Bleeding and haematoma | 2 | - | 1 | 2 | 5 | - | 1 | - | 1 | - |
| Bronchospasm | - | - | - | - | 1 | - | - | - | - | - |
| Nausea | - | - | 2 | - | - | - | - | - | - | - |
| Pseudoaneurysm | - | - | - | 1 | - | - | - | - | - | - |
| Vascular dissection | - | - | - | - | - | - | 1 | - | - | - |
| Vein rupture | 1 | - | - | - | - | - | - | - | 3 | 1 |
| Death** | - | - | - | - | - | - | 4 | - | 1 | - |
| **Not specified** | **0** | **-** | **0** | **0** | **0** | **0** | - | **18** | **12** | **0** |

*: exact number of individuals has not been specified

**: Death due to: AMI [21, 24] and cardiogenic shock [24]
