## Supplementary Table 5 for "Percutaneous rheolytic thrombectomy of thrombosed arteriovenous dialysis access using the AngioJet catheter- A Systematic Review"

| **First Author, Year** | **Follow-up duration** | **Success (%)** | | | **Patency rates (%)** | | |
| --- | --- | --- | --- | --- | --- | --- | --- |
|  |  | **Anatomic** | **Clinical** | **Technical** | **Primary** | **Assisted Primary** | **Secondary** |
| Adyin, 2020  [31] | 6 and 12 months | NR | NR | NR | 85.0, 78.0 | NR | NR |
| Drouven, 2020  [29] | 3, 6, 9 and 12 months | NR | AVF: 92.6  AVG: 92.0 | AVF: 92.6  AVG: 90.8 | AVF: 70.4, 51.9, 44.4, 40.7  AVG: 43.1, 29.2, 20.0, 10.8 | AVF: 85.2, 74.1, 63.0, 51.9  AVG: 61.5, 49.2, 33.9, 20.0 | AVF: 85.2, 74.1, 63.0, 51.9  AVG: 64.6, 58.5, 44.6, 35.4 |
| Lee, 2020  [28] | 3, 6 and 12  months | NR | 93.1 | 96.6 | 46.9, 39.7, 30.2 | NR | 82.5, 78.9, 75.3 |
| Ierardi, 2020  [30] | 3, 6, 12, 24  and > 24 months | NR | 91.7 | 95.0 | 98.0, 96.1, 72.5, 0.00, 0.00 | NR | 98.0, 98.0, 84.3, 31.4, 7.8 |
| Bermudez, 2016  [27] | 3, 6 and 12  months | NR | 86.0 | 93.0 | 52.0, 41.0, 23.0 | NR | 76.0, 68.0, 57.0 |
| Maleux, 2015  [26] | 3, 6, and 12  months | NR | 97.0 | 93.0 | 77.6, 62.4, 56.1 | 79.4, 69.9, 61.6 | 94.9, 94.9, 86.6 |
| Simoni, 2012  [24] | 0 and 3  months | NR | NR | AVF: 82.0  AVG: 98.0 | 0-month overall: 88.0  3-month AVF: 86.0  3-month AVG: 53.0 | NR | NR |
| Yang, 2012  [23] | 1, 6, 12  months | 77.0 | 76.0 | NR | 70.0, 45.0, 30.0 | NR | 76.0, 74.0, 74.0 |
| Litter, 2009  [21] | 0, 1, 3, 6, and  12 months | NR | NR | NR | Overall: 91.0, 71.0, 60.0, 37.0  AVF: 89.0, 67.0, 61.0, 34.0  AVG: 95.0, 80.0, 57.0, 43.0 | NR | NR, NR, 87.0, 77.0, 62.0 |
| Kakkos, 2008  [22] | 1, 6, 12, and 18 months | NR | 95.1 | 98.0 | 70.0, 40.0, 26.0, 18.0 | 72.4, 45.1, 30.0, 22.4 | NR |
