## Supplementary material for "Percutaneous rheolytic thrombectomy of thrombosed arteriovenous dialysis access using the AngioJet catheter- A Systematic Review": Figure 1

**549**	Articles identified through

database searching

**232** MEDLINE

**294** EMBASE

**23**	Cochrane

**102**	Excluded

**2**	Animal vein thrombosis models

**3**	Editorial

**10**	Only abstract available

**68**	Not relevant to AngioJet device

**2**	Same participants recruited

**17**	Single case report

**199**	Records

excluded on

title or abstract

**311**	Records screened

**238**	Duplicates removed

**10**	Studies included in the current systematic review

**112**	Full-text articles assessed for eligibility
